# Germline Mutation Analysis in Sporadic Breast Cancer Cases with Clinical Correlations

**DOI:** 10.1101/2021.03.29.21254525

**Authors:** Sadia Ajaz, Sani-e-Zehra Zaidi, Saleema Mehboob Ali, Aisha Siddiqa, Muhammad Ali Memon

**Author notes:** Corresponding Author: Sadia Ajaz, *Ph. D.

## Abstract

Demographics for breast cancers vary widely among nations. The frequency of germline mutations in breast cancers, which reflects the hereditary cases, has not been investigated adequately in Pakistani population. In the present case series, germ-line mutations in twenty-seven breast cancer candidate genes were investigated in eighty-four sporadic breast cancer patients along with the clinical correlations. The germ-line variants were also assessed in two healthy gender-matched controls. The most frequent parameters associated with hereditary cancer cases are age and ethnicity. Therefore, the clinico-pathological features were evaluated by descriptive analysis and Pearson *Χ*^2^ test (with significant p-value <0.05). The analyses were stratified on the basis of age (≤40 years vs >40 years) and ethnicity. The breast cancer gene panel assay was carried out by a genomic capture, massively parallel next generation sequencing assay on Illumina Hiseq2000 assay with 100bp read lengths. Copy number variations were determined by partially-mapped read algorithm. Once the mutation was identified, it was validated by Sanger sequencing. The ethnic analysis stratified on the basis of age showed that the frequency of breast cancer at young age (≤40 years) was higher in Sindhis (n=12/19; 64%) in contrast to patients in other ethnic groups. Majority of the patients had stage III (38.1%), grades II and III (46.4%), tumor size 2-5cm (54.8%), and invasive ductal carcinoma (81%). Overall, the analysis revealed germ-line mutations in 11.9% of the patients. The mutational spectrum was restricted to three genes: *BRCA1, BRCA2*, and *TP53*. The identified mutations consist of seven novel germ-line mutations, while three mutations have been reported previously. All the mutations are predicted to result in protein truncation. No mutations were identified in the remaining twenty-four candidate breast cancer genes. The present study provides the framework for the development of preventive and treatment strategies against breast cancers in Pakistani population.

## Introduction

Epidemiological studies have shown ethnic and geographic differences in breast cancer etiology. The increased susceptibility to breast cancers has been attributed to socio-economic, environmental, and genetic factors [1]. Hereditary breast cancers comprise a significant number. In the US population, these constitute 10-15% of the cases. There is paucity of data from low-to middle-income countries (LMIC) [2,3].

It has been estimated that half of all breast cancer cases occur in the 12% women who are at the maximum genetic risk [4]. *BRCA1* and *BRCA2* are high penetrant breast cancer genes. These have been especially associated with hereditary breast and/or ovarian cancers. The mutations in these genes are considered to increase the life-time risks of breast cancer by 82% [5,6]. Other highly penetrant but rare genes include *PTEN, TP53, CDH1*, and *STK11*. Moderate penetrance genes, which increase the risk for breast cancer by twofold, include genes involved in DNA repair such as *ATM, BRIP1* (*BACH1*), *CHEK2*, and *PALB2*. Still other genes are considered to confer a low but significant risk for breast cancers [7].

In Pakistani population, some studies have analyzed *BRCA1* and *BRCA2* mutations [8-15], mainly through conventional methodlogies. In case of other putative breast cancer susceptibility genes, scarce or no data is available from this region [16-20]. The introduction of next generation sequencing (NGS) including multi-gene testing necessitates re-assessment of the available information and generation of missing data.

Pakistani population comprises distinct ethnic groups. These include Sindhis, Balochis, Brahui, Makrani, and Parsis from Southern Pakistan. The other ethnicities Punjabis, Pathans, Hindko, Hazara, Kalash, Kashmiri and Burusho are from Northern Pakistan [20, 21]. The data for these genetically distinct ethnic groups has not been incorporated in the few relevant regional-based publications on the breast cancers. The location of present study is a metropolitan city, situated in Southern Pakistan. The population comprises multiple ethnic communities from all over Pakistan. In addition, a self-defined Urdu-speaking ethnicity, comprising immigrants from India, is also a major group residing in the city.

The present study investigates molecular epidemiology of breast cancers from Southern Pakistan. It is the first such report from this region. The study investigates the genetic contribution to breast cancers by next-generation sequencing. A panel of twenty-seven breast cancer-associated candidate genes, has been analysed in breast cancer patients belonging to genetically distinct Pakistani ethnic groups.

## Materials and methods

### Patients

Participants in the present study included 82 females and 02 males from Southern Pakistan. In total, 84 diagnosed cases of breast cancer were included in the study. The participants visited a tertiary care hospital: the Atomic Energy Medical Centre (AEMC), Jinnah Post-Graduate Medical Centre (JPMC), Karachi, Pakistan, from July, 2016 – July, 2017. The patients were treated for primary invasive breast cancer post-mastectomy.

At the preliminary stage, all the clinically diagnosed primary breast cancer cases were included regardless of the age and/or family history. All the participants signed an informed consent form. Independent ethical review boards of the participating institutions approved the protocol.

### Demographic and Clinico-pathological Information

Patients were interviewed about their family history of cancers (breast cancer and/or any other cancer), ethnicity, age at menarche and menopause (if applicable), and gynae & obs history. The medical records were reviewed for breast cancer diagnosis, staging, grading, and tumour size.

### Genomics

The participants contributed 5-8ml of blood samples for DNA extraction. Germ-line DNA was extracted from the patient’s WBCs by standard phenol-chloroform method [22]. DNA was quantified spectrophotometrically (Beckman Coulter™, DU^®^ 530). Sufficient DNA was available for 84 subjects. BROCA [7], a targeted capture and multiplexed massively parallel sequencing gene panel assay was performed. It enables detection of all types of mutations for candidate and established breast cancer genes. Twenty-seven genes, which are highly associated with breast cancers, were investigated in the project: *BRCA1, BRCA2, TP53, ATR, BARD1, BRIP1, FAM175A, FANCM, GEN1, MRE11A, NBN, RAD51B, RAD51C, RAD51D, RECQL, RINT1, SLX4, BAP1, PALB2, PTEN, STK11, XRCC2, ATM, CHEK1, CHEK2, CDH1*, and *CTNNA1*.

### Validation of Mutations

After the NGS investigations, the identified mutations were validated by Sanger sequencing. Previously published protocols were used for the amplification of exons [23-25] followed by standard method for Sanger sequencing.

### Statistical and Bio-informatic Analysis

Data were entered, encoded and analysed using SPSS, version 17.0 (IBM™, USA). Breast cancer cases in the present study were grouped into three categories: age at sampling (≤40years vs >40 years); receptor (estrogen, progesterone, and HER2/Neu) status; and ethnicities. Descriptive analysis was carried out for the evaluation of demographics and clinico-pathological features. Groups were compared by Pearson χ^2^ test of independence for the clinico-pathological features: tumour size, grade, and stage. The p-values <0.05 were considered to be statistically significant.

The mutations were compared against BIC [25] and ExAC [26] databases for novelty. The consequences of identified mutations were analysed as described previously [27].

## Results

### Demographic Analysis

Total study included 84 breast cancer patients, with a diagnosis of primary breast cancer. Patients’ demographics are shown in Table 1. In the present cohort, majority of the breast cancer cases belonged to Urdu-speaking (25%) and Sindhi (24%) ethnicities. The frequency of breast-cancer cases among young patients (≤ 40 years) was higher in Hindko (n=4; 75%) and Sindhi (n=19; 64%) ethnicities, in contrast to other ethnic groups. Among other ethnic groups, the numbers of breast cancer cases in older patients (i.e. > 40 years) exceeded those who were ≤ 40 years.

**Table 1.** Patients’ demographics.

| **Sr. No.** | **Characteristic** | **Total** | **Patients' Age ≤ 40 years** | **Patients' Age > 40 years** |
| --- | --- | --- | --- | --- |
| 1. | Total unrelated patients | 84 | 35 (42%) | 49 (58%) |
|  | Female patients | 82 | 35 (43%) | 47 (57%) |
|  | Male patients | 02 | 00 (0%) | 02 (100%) |
| 2. | Mean Age<sub>samp</sub> (years) | 44 (20-70) | 35 (20 – 40) | 51 (41 – 70) |
| 4. | Ethnicity* |  |  |  |
|  | Baloch | 08 | 02 (25%) | 06 (75%) |
|  | Hindko | 04 | 02 (50%) | 02 (50%) |
|  | Punjabi | 05 | 02 (40%) | 03 (60%) |
|  | Pathan | 9 | 03 (33%) | 06 (67%) |
|  | Sindhi | 19 | 12 (63%) | 07 (37%) |
|  | Urdu-Speaking | 19 | 09 (47%) | 10 (53%) |
|  | Others | 11 | 02 (18%) | 09 (82%) |

\*ethnicities were unknown for 09 patients.

### Clinicopathological Evaluation

Pathology records were sought for the patients. Data were available for 70.9%, 95%, and 82.5% patients in case of tumour stage, grade and hormonal status, respectively. Among patients with available pathology data, the distribution of tumor stage was 5% Stage I, 33% Stage II, 56% Stage III, and 6% Stage IV. The distribution of tumour grade was 1% Grade I, 48% Grade II, 49% Grade III, and 2% for Grade IV. Overall, the distribution among stages and grades varied significantly (p<0.01). In case of tumours with available hormone profiles, 26% were triple negative (TNBC). Supplementary table 1 lists the available clinicopathological information.

### Novel Germ-line Mutations

A total of 84 samples were analyzed based upon sufficient DNA quantity. Genomic analysis of known breast cancer genes revealed that 11.9% (10/84) patients carried an unambiguously pathogenic germline mutation in three genes: *BRCA1, BRCA2*, and *TP53* (Table 2). Novel germ-line mutations were identified in seven patients (3 in *BRCA1*, 3 in *BRCA2*, and 1 in *TP53*) (Table 2 and Supplementary Figure).

**Table 2.** Summary of germline mutations identified in *BRCA1, BRCA2*, and *TP53* genes in Pakistani breast cancer patients.

| Sr<br>·<br>N | ID | Histology<sup>¶</sup> | Sex | Age<br>Group | Chrom<br>osome | Position<br>Start hg 37 | Gene | c. DNA | Protein | Novel*<br>Mutation |
| --- | --- | --- | --- | --- | --- | --- | --- | --- | --- | --- |
| 1. | 09 | pBC (IDC) | F | ≤40 | 13 | 32,914,299 | *BRCA2* | c.5807delTGTC (ex11) | 1936fs | yes |
| 2. | 38 | pBC (IDC) | F | >40 | 17 | 41,276,045 | *BRCA1* | c.68_69del (ex2) | 23fs | no |
| 3. | 45 | pBC (IDC) | F | ≤40 | 17 | 41,244,237 | *BRCA1* | c.3311delA (ex11) | K1104fs | yes |
| 4. | 47 | pBC (IDC) | F | ≤40 | 17 | 41,253,030 | *BRCA1* | del exons 5-7 | K45fs | yes |
| 5. | 63 | pBC (IDC) | F | ≤40 | 17 | 7,578,212 | *TP53* | c.637C>T (ex6) | R213X | no |
| 6. | 71 | pBC (IDC) | F | >40 | 13 | 32,954,023 | *BRCA2* | c.9090delA (ex23) | T3030fs | yes (germline) |
| 7. | 73 | pBC (IDC) | F | ≤40 | 17 | 41,197,784 | *BRCA1* | c.5503C>T (ex24) | R1835X | no |
| 8. | 84 | pBC (IDC+DCIS) | F | ≤40 | 17 | 41,245,795 | *BRCA1* | c.1753G>T (ex11) | E585X | yes |
| 9. | 88 | pBC (IDC) | F | >40 | 13 | 32,914,134 | *BRCA2* | c.5642delAATC (ex11) | 1881fs | yes |
| 10 | 93 | pBC (IDC) | F | >40 | 17 | 7,579,415 | *TP53* | c.272G>A (ex4) | W91X | yes (germline) |

<sup>¶</sup>pBC: primary Breast Cancer; IDC: Invasive Ductal Carcinoma; DCIS: Ductal Carcinoma *in situ*).
\*Novelty was identified by comparing with BIC, and ExAC databases.

### Burden of Mutations in Breast Cancer Genes

Genomic analysis of known breast cancer genes showed that 15.3% (6/39) of patients with age ≤ 40 years, whereas 8.5% (4/47) of patients with age > 40 years carried a definitive pathogenic germline mutation in three identified genes (Table 3).

**Table 3.** Distribution of germ-line mutations in breast cancer patients. Data has been stratified on the basis of age, hormone receptor status, and ethnicities.

| Sr. No. | Characteristic | Total Carriers | Frequency of Mutation Carriers |  |  |
| --- | --- | --- | --- | --- | --- |
|  |  |  | *Any Gene* | *BRCA1/2* | *TP53* |
| 1 | **Total patients with mutations** | 10 | 0.12 (10/84) | 0.1 (08/84) | 0.03 (02/84) |
| 2 | **Age at sampling (Age<sub>samp</sub>)** |  |  |  |  |
|  | Age<sub>samp</sub> ≤ 40 years | 6 | 0.15 (6/39) | 0.13 (5/39) | 0.03 (1/39) |
|  | Age<sub>samp</sub> > 40 years, positive FH | 4 | 0.08 (4/51) | 0.06 (3/51) | 0.02 (1/51) |
| 3 | **Tumour hormone receptors** |  |  |  |  |
|  | TNBC | 4 | 0.23 (4/17) | 0.23 (4/17) | 0 |
|  | Non TNBC | 3 | 0.06 (3/49) | 0.02 (1/49) | 0.04 (2/49) |
|  | Unknown | 3 | 0.3 | 0.2 | 0.1 |
| 4 | **Ethnicity\*** |  |  |  |  |
|  | Baloch | 0 | 0 | 0 | 0 |
|  | Hindko | 2 | 0.5 (2/4) | 0.5 (2/4) | 0 |
|  | Punjabi | 1 | 0.13 (1/8) | 0.13 (1/8) | 0 |
|  | Pathan | 0 | 0 | 0 | 0 |
|  | Sindhi | 2 | 0.11 (2/19) | 0.06 (1/19) | 0.06 (1/19) |
|  | Urdu-Speaking | 3 | 0.15 (3/20) | 0.15 (3/20) | 0 |
|  | Others | 0 | 0 | 0 | 0 |
|  | Unknown | 2 | 0.18 (2/17) | 0.18 (2/17) | 0 |

Among younger patients, 13% (5/39) carried a damaging mutation in *BRCA1* and *BRCA2* while 3% (1/39) carried a germ-line mutation in *TP53*. In the older patients, 5.8% (3/51) harboured a germ-line mutation in *BRCA1* and *BRCA 2*, whereas the frequency of germ-line mutation in *TP53* was 2% (1/51). Stratified analysis showed that the highest frequency of germ-line mutations in the investigated genes was in Hindko group (50%), followed by Urdu-speaking (15%), Punjabi (13%), and Sindhi (11%) ethnicities.

Among patients with known tumour hormone receptor status, 23% (4/17) TNBC patients carried a pathogenic germline mutation in *BRCA1*/*2* genes (2 in *BRCA1* and 2 in *BRCA2*).

Each identified mutation was present in only one family and no recurrent mutation was found in the present cohort.

No pathogenic germ-line variants were identified in the DNA from control samples.

## Discussion

Breast cancer is the most frequently diagnosed malignancy among women and the leading cause of cancer-related mortality in developing countries [28]. It is estimated that globally 1 in 6 women is diagnosed with breast cancer and 1 in 8 women has invasive form of the cancer. However, substantial differences have been observed in breast cancer indices across different populations [29]. The average age in Caucasians is 63 years. In the present study we report an average of 44.4 years, whereas from the same region it is reported in the range of 50-53 years from India, and 46-49 years from Iran, respectively [30-33].

Majority of familial aggregation in breast cancers is unexplained. Environmental factors are unlikely to explain the residual familial clustering [4]. Among Caucasians, with the increasing application of next generation sequencing in the clinical setting, an upward trend (26%) in reports of hereditary breast and/or ovarian cancers is observed [5]. A number of large scale studies report germ-line mutational frequencies ranging from 9%-26% in critical breast cancer genes [34-37].

In case of non-Caucasian females, there is paucity of data regarding molecular basis of breast cancers. To the best of our knowledge the present study is the first report of a twenty-seven breast cancer gene panel analysis in a South-Asian population. The molecular investigation includes high and moderate penetrance genes [7]. Here we report germ-line mutations in three high penetrance genes: *BRCA1, BRCA2*, and *TP53* in breast cancer patients from this population. The identified mutations consist of seven novel germ-line mutations, while three mutations have been reported previously. The location of inherited germ-line mutations the genes is shown in figure 1.

**Figure 1.**
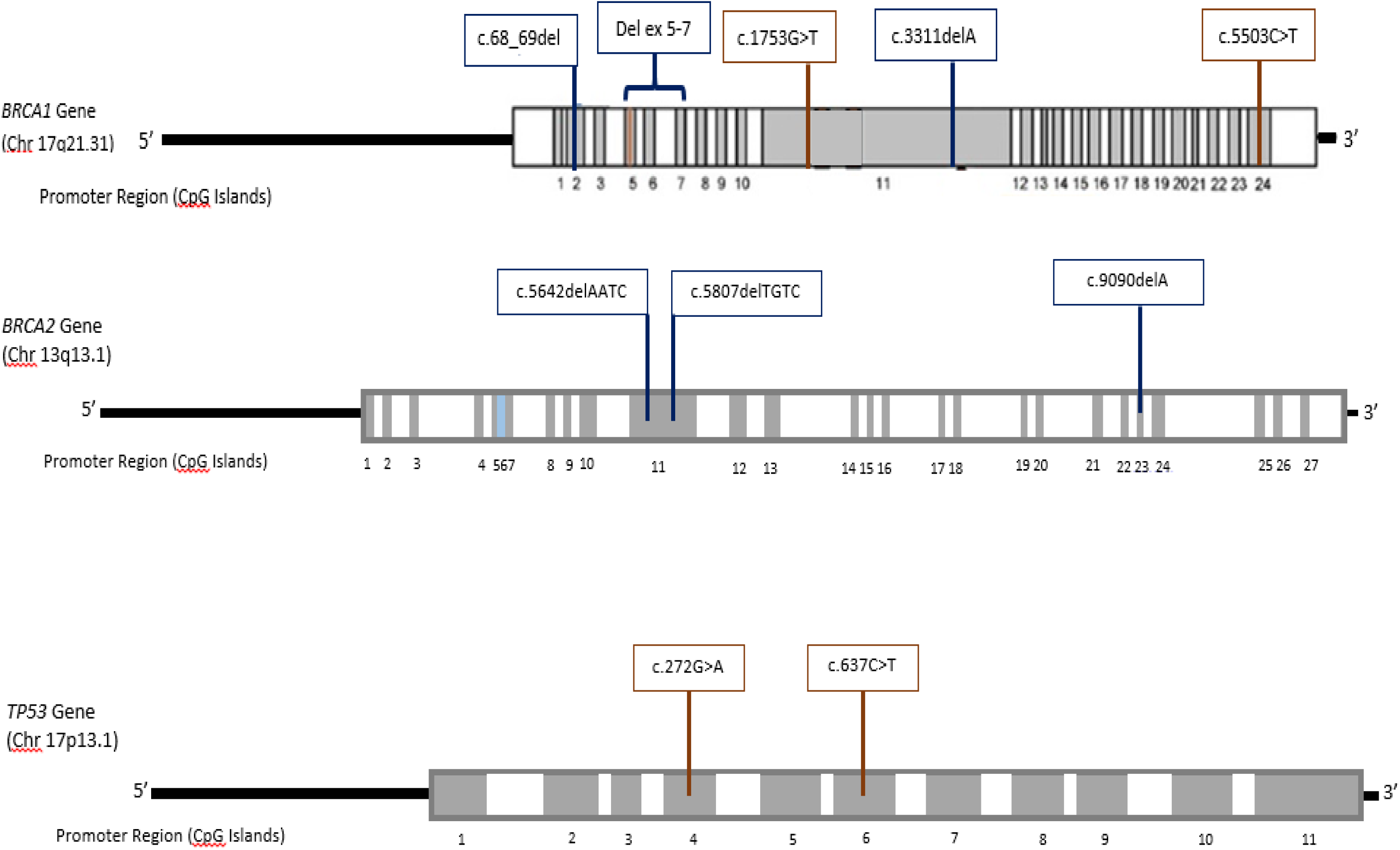
Exon-specific distribution of identified germ-line mutations in *BRCA1, BRCA2*, and *TP53* genes.

The identified mutations were heterozygous. Bi-allelic *BRCA1* mutations are likely to be lethal at the embryonic stage, while such mutations in *BRCA2* lead to Fanconi anemia type D1, with increased risk of childhood cancer [9, 38]. Germline mutations in *TP53* are associated with Li-Fraumeni syndrome. In the present cohort, no syndromic cases were identified.

The founder mutations are expected in consanguineous populations like the present one. Although we did not find any recurrent mutation in the present cohort, comparison with previously published reports indicated that 185delAG may be a founder mutation [8-15].

Germline mutations were not identified in the rest of reported high penetrant genes including *PTEN, CDH1*, and *STK11* [39-43]

Moderate penetrance genes include additional DNA repair genes. These are *CHEK2* (, *BRIP1* (*BACH1*), *ATM, PALB2* [44-47]. These interact with *BRCA1* and/or *BRCA2*. The mutations in these genes result in two-fold increase in breast cancer risk. In the present study, no germ-line mutation was identified in these genes.

Similarly, no germline mutations were identified in other candidate genes *ATR, BARD1, FAM175A, FANCM, GEN1, MRE11A, NBN, RAD51B, RAD51C, RAD51D, RECQL, RINT1, SLX4, BAP1, XRCC2, CHEK1*, and *CTNNA1* [48-69]

Interestingly, all the identified mutations are nonsense mutations and predicted to result in protein truncation. This corroborates the data for *BRCA1* and *BRCA2*, but not for *TP53* from India. Among South-Asian populations, the germ-line mutation rate (11.9%) in the present study is three folds less than reported for India [30]. In contrast to the reported observations that younger patients are likely to be the carriers of germ-line mutation, we report a lower frequency of such mutations as compared to the frequency of such mutations from India (36%). Interestingly, the average age of the patients in their study [70] is higher as compared to the present report (50 vs 44 years).

Most studies on *BRCA1* and *BRCA2* mutations from Asia report a higher frequency for *BRCA2* mutations than *BRCA1*, the exceptions being Pakistan and India [71]. The pattern is also observed in the present study.

The present study is also the first report of higher frequency of younger breast cancer patients belonging to Hindko and Sindhi ethnicities as compared to the other ethnicities in the region. As the investigated genes do not account for all such cases, it is possible that as yet unidentified gene(s) may be involved in these ethnic groups. It is pertinent to mention that the present day Pakistan consists of more than 12 distinct ethnic and linguistic groups [72, 73]

In conclusion, the present study while providing a framework for the investigation of genetic basis of breast cancers for cost-effective screening and management, raises many questions. The foremost is: as the germline mutations account for only 12% of the breast cancer cases, which other factors (genetic and/or environmental) are involved in the observed high incidence of breast cancers? It is expected that building on the present findings, a scientifically-focused approach may be developed for breast cancer research in a resource-limited setting.

## Supporting information

STROBE CHECKLIST

## Data Availability

Data available on reasonable request

## Acknowledgements

The authors wish to thank Prof. MC King and Prof. Tom Walsh for BROCA analysis, ICCBS for core facilities, and AEMC, JPMC staff for their co-operation. The authors are especially grateful to the participants in the study.

## Compliance with ethical standards

### Availability of data and material

The data and information which has been analysed in the present report are available from the corresponding author upon request.

### Ethical approval

The study was conducted in accordance with the Declaration of Helsinki [21]. The project was approved by the ethical review committees (ERCs) of the participating institutions: the independent ERC, International Center for Chemical and Biological Sciences (ICCBS), University of Karachi, Karachi, Pakistan [ICCBS/IEC-016-BS/HT-2016/Protocol/1.0], and the Atomic Energy Medical Centre (AEMC), Jinnah Postgraduate Medical Centre (JPMC), Karachi, Pakistan [Admin-3(257)/2016].

### Informed Consent

All the samples were collected after obtaining written informed consent from each participant.

## TABLES AND FIGURES

**Supplementary Table 1.** Clinicopathological Characteristics of the Breast Cancer Samples for the.

| *Sr. No.* | *Tumour characteristic* | *Total number of samples* | *Value/ numbers* |
| --- | --- | --- | --- |
| **1.** | **Tumour Size** | **76** | **<2cm : 10 (11.9%)**<br>**2-5cm : 46 (54.8%)**<br>**>5cm : 20 (23.8%)** |
| **2.** | **Tumour Stage** | **58** | **T<sub>1</sub>: 03 (03.6%)**<br>**T<sub>2</sub>: 19 (22.6%)**<br>**T<sub>3</sub>: 32 (38.1%)**<br>**T<sub>4</sub>: 04 (4.8%)** |
| **3.** | **Tumour Grade** | **80** | **G1: 01 (1.2%)**<br>**G2: 39 (46.4%)**<br>**G3: 39 (46.4%)**<br>**G4: 1 (1.2%)** |
| **4.** | **Histopathology\*** | **84** | **IDC: 68 (81%)**<br>**ILC: 01 (1.2%)**<br>**DCIS + IDC: 11 (13.1%)**<br>**Others: 04 (4.80%)** |

\*IDC: Invasive Ductal Carcinoma; DCIS: Ductal Carcinoma Insitu; In situ; ILC: Invasive Lobular Carcinoma

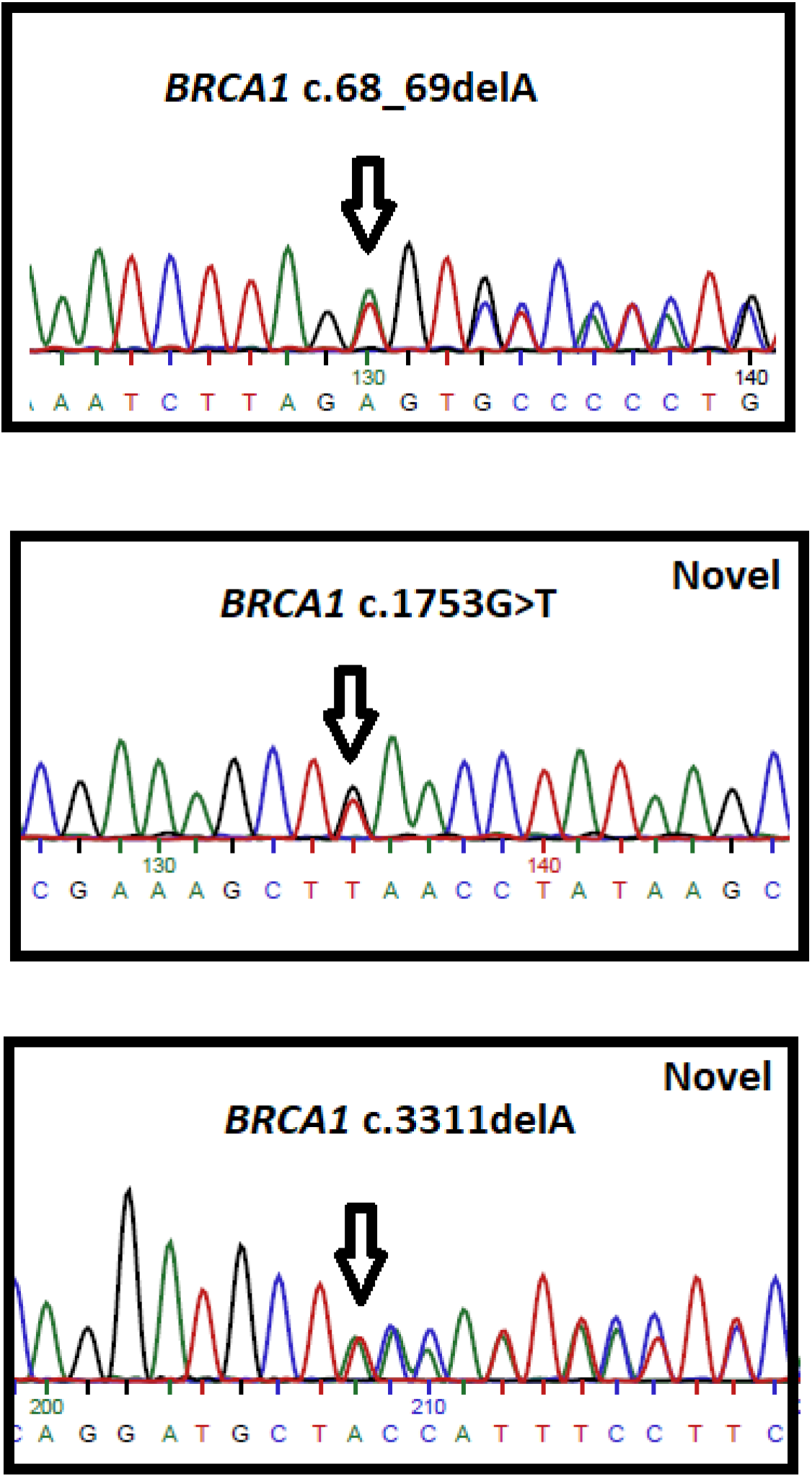

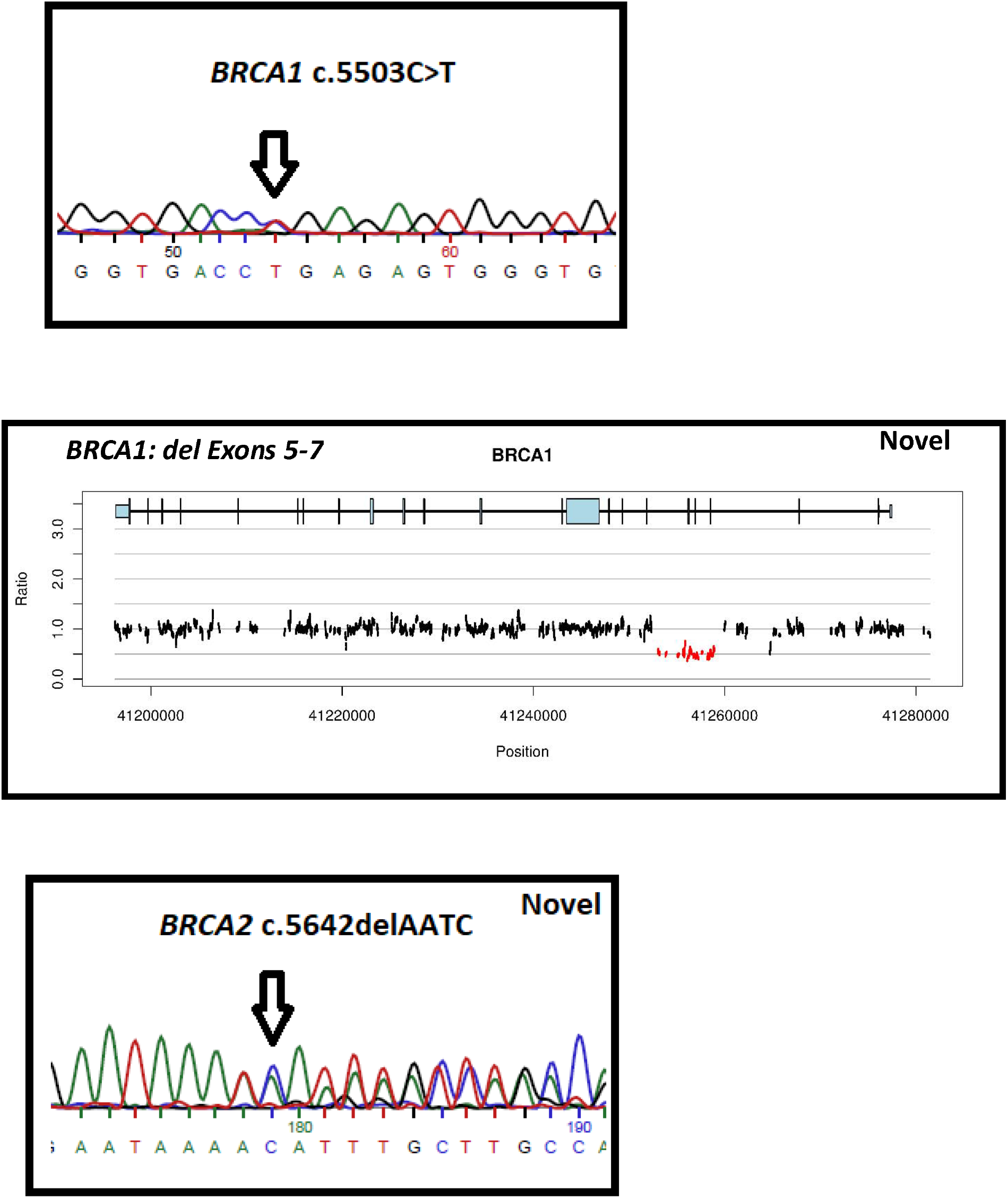

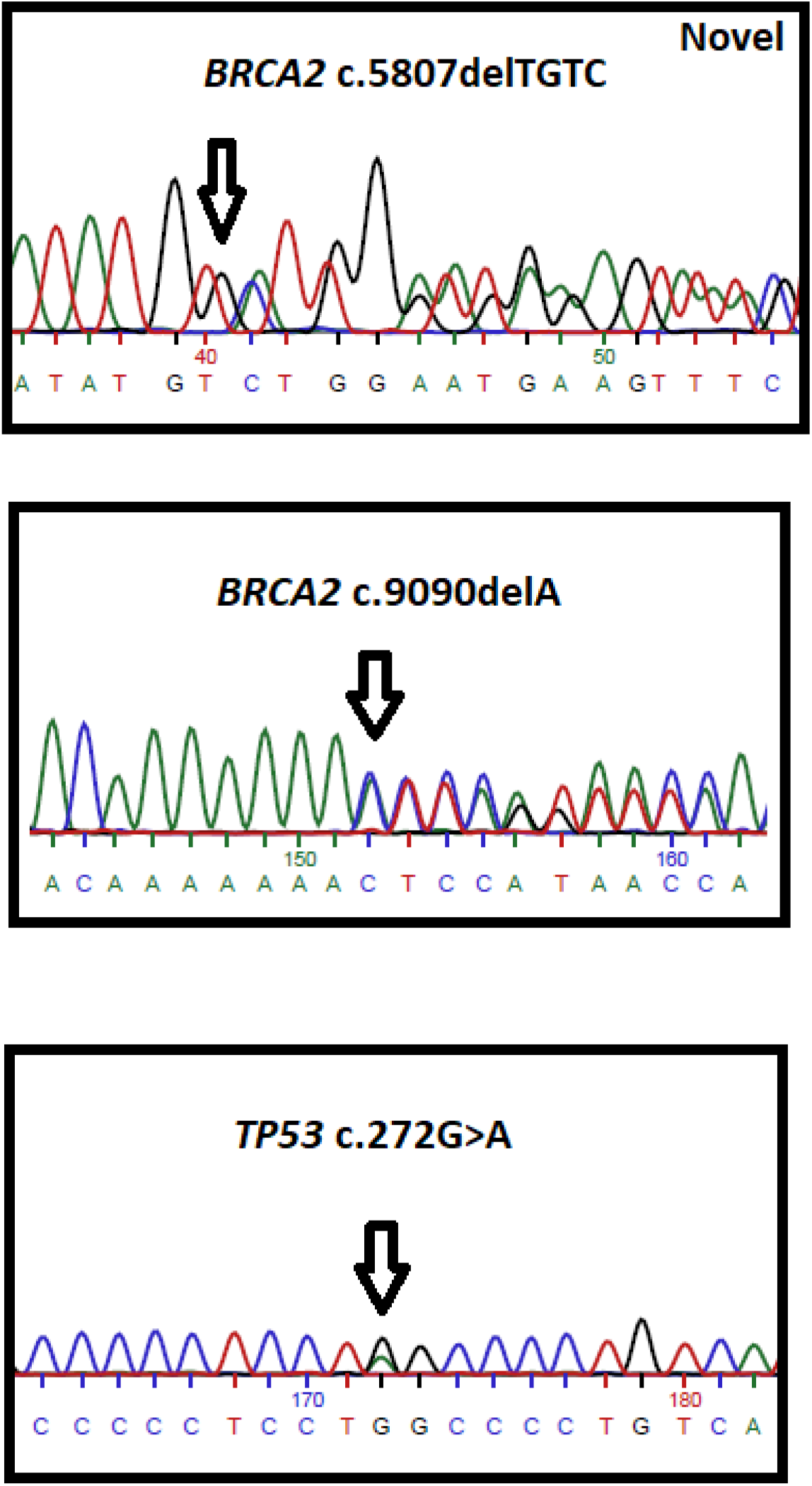

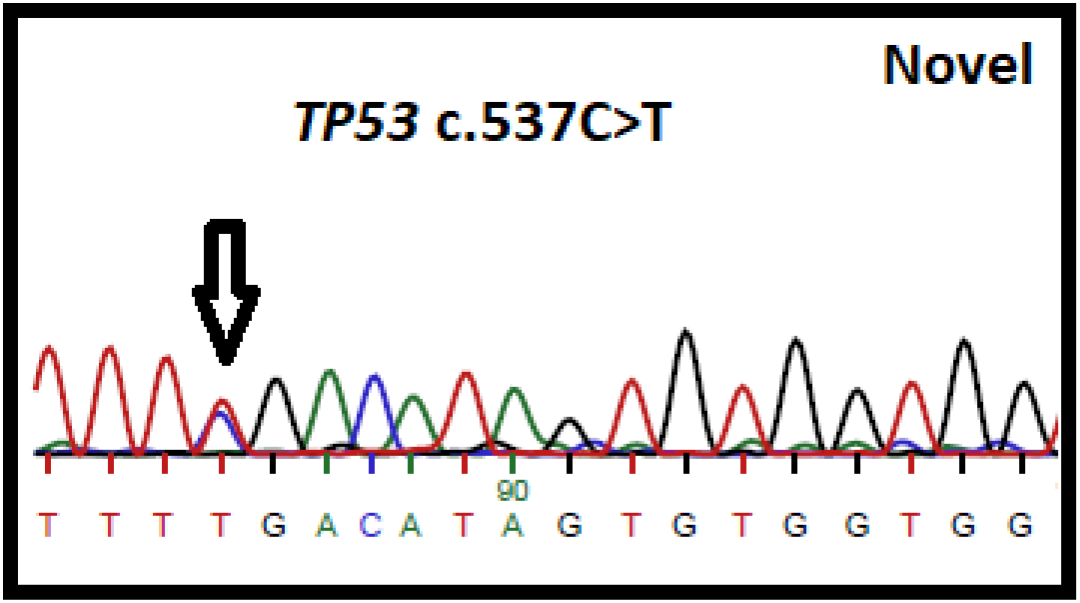

## References

1. Zeeshan S, Ali B, Ahmad K, Chagpar AB et al (2019) Clinicopathological Featuresof Young Versus Older Patients With Breast Cancer at a Single Pakistani Institution and a Comparison With a National US Database. J Glob Oncol 5:1–6. doi: 10.1200/JGO.18.00208

2. Akarolo-Anthony SN, Ogundiran TO, Adebamowo CA (2010) Emerging breast cancer epidemic: evidence from Africa. Breast Cancer Res 20;12 Suppl 4:S8.doi: 10.1186/bcr2737

3. Jazayeri SB, Saadat S, Ramezani R et al (2015) Incidence of primary breast cancer in Iran: Ten-year national cancer registry data report. Cancer Epidemiol 39(4):519–27. doi: 10.1016/j.canep.2015.04.016. Epub 2015 Jun 10.

4. El Saghir NS, Khalil MK, Eid T et al (2007) Trends in epidemiology and management of breast cancer in developing Arab countries: a literature and registry analysis. Int J Surg 5(4):225-33. Epub 2006 Aug 24.

5. Antoniou AC, Easton DF (2006) Models of genetic susceptibility to breast cancer. Oncogene. 25(43):5898–905.

6. Shiovitz S, Korde LA (2015) Genetics of breast cancer: a topic in evolution. Ann Oncol. 2015 Jul;26(7):1291–9. doi: 10.1093/annonc/mdv022. Epub 2015 Jan 20.

7. Walsh T, Mandell JB, Norquist BM et al (2017) Genetic Predisposition to Breast Cancer Due to Mutations Other Than BRCA1 and BRCA2 Founder Alleles Among Ashkenazi Jewish Women. JAMA Oncol. 1;3(12):1647–1653. doi: 10.1001/jamaoncol.2017.1996.

8. Liede A, Malik IA, Aziz Z et al (2002) Contribution of BRCA1 and BRCA2 mutations to breast and ovarian cancer in Pakistan. Am J Hum Genet. 2002 Sep;71(3):595-606. Epub 2002 Aug 13.

9. Moatter T, Aban M, Khan S et al (2011) BRCA1 status in Pakistani breast cancer patients with moderate family history. J Coll Physicians Surg Pak. 21(11):680–4. doi: 11.2011/JCPSP.680684.

10. Ahmad J, Le Calvez-Kelm F, Daud S et al (2012) Detection of BRCA1/2 mutations in breast cancer patients from Thailand and Pakistan. Clin Genet. 82(6):594–8. doi: 10.1111/j.1399-0004.2012.01869.x. Epub 2012 Apr 8.

11. Rashid MU, Zaidi A, Torres D et al (2006) Prevalence of BRCA1 and BRCA2 mutations in Pakistani breast and ovarian cancer patients. Int J Cancer. 119(12):2832–9.

12. Aziz F, Fatima W, Mahmood S et al (2016) Screening for Del 185 AG and 4627C>A BRCA1 Mutations in Breast Cancer Patients from Lahore, Pakistan. Asian Pac JCancer Prev. 17(4):1725–7.

13. Rashid MU, Muhammad N, Amin A et al (2017) Contribution of BRCA1 large genomic rearrangements to early-onset and familial breast/ovarian cancer in Pakistan. Breast Cancer Res Treat. 2017 Jan;161(2):191–201. doi:10.1007/s10549-016-4044-0. Epub 2016 Nov 8.

14. Rashid MU, Muhammad N, Bajwa S et al (2016) High prevalence and predominance of BRCA1 germline mutations in Pakistani triple-negative breast cancer patients. BMC Cancer. 23;16(1):673. doi: 10.1186/s12885-016-2698-y.

15. Torres D, Bermejo JL, Rashid MU et al (2017) Prevalence and Penetrance of BRCA1 and BRCA2 Germline Mutations in Colombian Breast Cancer Patients. Sci Rep. 2017 Jul 5;7(1):4713. doi:10.1038/s41598-017-05056-y.

16. Baig RM, Mahjabeen I, Sabir M et al (2011) Genetic changes in the PTEN gene and their association with breast cancer in Pakistan. Asian Pac J Cancer Prev. 12(10):2773–8.

17. Baloch AH, Khosa AN, Bangulzai N et al (2016) Novel Nonsense Variants c.58C>T (p.Q20X) and c.256G>T (p.E85X) in the CHEK2 Gene Identified in Breast Cancer Patients from Balochistan. Asian Pac J Cancer Prev. 2016;17(7):3623–6.

18. Rashid MU, Khan FA, Muhammad N et al (2018) Prevalence of PALB2 Germline Mutations in Early-onset and Familial Breast/Ovarian Cancer Patients from Pakistan. Cancer Res Treat. 2018 Oct 11. doi: 10.4143/crt.2018.356.

19. Rashid MU, Muhammad N, Faisal S (2014) Deleterious RAD51C germline mutations rarely predispose to breast and ovarian cancer in Pakistan.Breast Cancer Res Treat. 2014 Jun;145(3):775–84. doi: 10.1007/s10549-014-2972-0.

20. Khaliq S, Hameed A, Khaliq T (2000) P53 mutations, polymorphisms, and haplotypes in Pakistani ethnic groups and breast cancer patients. Genet Test. 2000;4(1):23–9.

21. Qamar R, Ayub Q, Khaliq S (1999) African and Levantine origins of Pakistani YAP+ Y chromosomes. Hum Biol. 71(5):745–55.

22. Sambrook, J. and D.W. Russell, Molecular cloning: a laboratory manual. 2001. 2001, Cold Spring Harbor Laboratory Press, Cold Spring Harbor, New York.

23. Friedman LS, Ostermeyer EA, Szabo CI et al (1994) Confirmation of BRCA1 by analysis of germline mutations linked to breast and ovarian cancer in ten families. Nat Genet. 1994 Dec;8(4):399–404.

24. Friedman LS, Gayther SA, Kurosaki T et al (1997) Mutation analysis of BRCA1 and BRCA2 in a male breast cancer population. Am J Hum Genet.60(2):313–9.

25. Olivier M, Eeles R, Hollstein M et al (2002) The IARC TP53 database: new online mutation analysis and recommendations to users. Hum Mutat.19(6):607–14.

26. BIC: https://research.nhgri.nih.gov/bic/ 27.

27. ExAC: http://exac.broadinstitute.org/

28. Ferlay J E.M., Lam F, et al (2018) Global Cancer Observatory: Cancer Today. Lyon, France: International Agency for Research on Cancer. Available from: https://gco.iarc.fr/today, accessed [24 December 2018]. 2018.

29. Bray F, Ferlay J, Laversanne M (2015) Cancer Incidence in Five Continents: Inclusion criteria, highlights from Volume X and the global status of cancer registration. Int J Cancer. 137(9):2060–71. doi: 10.1002/ijc.29670.

30. Mannan AU, Singh J, Lakshmikeshava R (2016) Detection of high frequency of mutations in a breast and/or ovarian cancer cohort: implications of embracing a multi-gene panel in molecular diagnosis in India. J Hum Genet. 61(6):515–22. doi: 10.1038/jhg.2016.4.

31. Howlader N, Noone AM, Krapcho M et al (2017) SEER Cancer Statistics Review, 1975–2014. Bethesda, MD: National Cancer Institute.

33. Ahmadi AS, Mahdipour L, Payandeh M et al (2015) Epidemiology, pathology and histochemistry features in women with breast cancer. Am J Cancer Prev. 3: p. 54–7.

34. Desmond A, Kurian AW, Gabree M et al (2015) Clinical Actionability of Multigene Panel Testing for Hereditary Breast and Ovarian Cancer Risk Assessment. JAMA Oncol. 1(7):943–51. doi: 10.1001/jamaoncol.2015.2690.

35. Buys SS, Sandbach JF, Gammon A et al (2017) A study of over 35,000 women with breast cancer tested with a 25-gene panel of hereditary cancer genes. Cancer. 123(10):1721–1730. doi: 10.1002/cncr.30498.

36. Eliade M, Skrzypski J, Baurand A et al (2017) The transfer of multigene panel testing for hereditary breast and ovarian cancer to healthcare: What are the implications for the management of patients and families? Oncotarget. 2017 Jan 10;8(2):1957–1971. doi: 10.18632/oncotarget.12699.

37. Castéra L, Krieger S, Rousselin A et al (2014) Next-generation sequencing for the diagnosis of hereditary breast and ovarian cancer using genomic capture targeting multiple candidate genes. Eur J Hum Genet. 22(11):1305–13. doi: 10.1038/ejhg.2014.16.

38. Stratton, M.R. and N. Rahman (2008) The emerging landscape of breast cancer susceptibility. Nature genetics 40(1): p. 17.

39. FitzGerald, M.G., et al., Germline mutations in PTEN are an infrequent cause of genetic predisposition to breast cancer. Oncogene, 1998. 17(6): p. 727.

40. Tan, M.-H., et al., Lifetime cancer risks in individuals with germline PTEN mutations. Clinical Cancer Research, 2012. 18(2): p. 400–407.

41. Pharoah, P., P. Guilford, and C. Caldas, International Gastric Cancer Linkage C. Incidence of gastric cancer and breast cancer in CDH1 (E-cadherin) mutation carriers from hereditary diffuse gastric cancer families. Gastroenterology, 2001. 121(6): p. 1348–1353.

42. Lim, W., et al., Relative frequency and morphology of cancers in STK11 mutation carriers1. Gastroenterology, 2004. 126(7): p. 1788–1794.

43. Boardman, L.A., et al., Increased risk for cancer in patients with the Peutz-Jeghers syndrome. Annals of internal medicine, 1998. 128(11): p. 896–899.

44. Meijers-Heijboer, H., et al., Low-penetrance susceptibility to breast cancer due to CHEK2(*)1100delC in noncarriers of BRCA1 or BRCA2 mutations. Nat Genet, 2002. 31(1): p. 55–9.

45. Seal, S., et al., Truncating mutations in the Fanconi anemia J gene BRIP1 are low-penetrance breast cancer susceptibility alleles. Nature genetics, 2006. 38(11): p. 1239.

46. Renwick, A., et al., ATM mutations that cause ataxia-telangiectasia are breast cancer susceptibility alleles. Nature genetics, 2006. 38(8): p. 873.

47. Rahman, N., et al., PALB2, which encodes a BRCA2-interacting protein, is a breast cancer susceptibility gene. Nature genetics, 2007. 39(2): p. 165.

48. Tanaka, A., et al., Germline mutation in ATR in autosomal-dominant oropharyngeal cancer syndrome. The American Journal of Human Genetics, 2012. 90(3): p. 511–517.

49. Ratajska, M., et al., Cancer predisposing BARD1 mutations in breast-ovarian cancer families. Breast Cancer Res Treat, 2012. 131(1): p. 89–97.

50. Solyom, S., et al., Breast cancer-associated Abraxas mutation disrupts nuclear localization and DNA damage response functions. Sci Transl Med, 2012. 4(122): p. 122ra23.

51. Kiiski, J.I., et al., Exome sequencing identifies FANCM as a susceptibility gene for triple-negative breast cancer. Proc Natl Acad Sci U S A, 2014. 111(42): p. 15172–7.

52. Turnbull, C., et al., Mutation and association analysis of GEN1 in breast cancer susceptibility. Breast Cancer Res Treat, 2010. 124(1): p. 283–8.

53. Bartkova, J., et al., Aberrations of the MRE11-RAD50-NBS1 DNA damage sensor complex in human breast cancer: MRE11 as a candidate familial cancer-predisposing gene. Mol Oncol, 2008. 2(4): p. 296–316.

54. Stewart, G.S., et al., The DNA double-strand break repair gene hMRE11 is mutated in individuals with an ataxia-telangiectasia-like disorder. Cell, 1999. 99(6): p. 577–87.

55. Madanikia, S.A., et al., Increased risk of breast cancer in women with NF1. Am J Med Genet A, 2012. 158a(12): p. 3056–60.

56. Golmard, L., et al., Germline mutation in the RAD51B gene confers predisposition to breast cancer. BMC Cancer, 2013. 13: p. 484.

57. Walsh, T., et al., Mutations in 12 genes for inherited ovarian, fallopian tube, and peritoneal carcinoma identified by massively parallel sequencing. Proc Natl Acad Sci U S A, 2011. 108(44): p. 18032–7.

58. Loveday, C., et al., Germline RAD51C mutations confer susceptibility to ovarian cancer. Nat Genet, 2012. 44(5): p. 475-6; author reply 476.

59. Loveday, C., et al., Germline mutations in RAD51D confer susceptibility to ovarian cancer. Nat Genet, 2011. 43(9): p. 879–882.

60. Shah, S., et al., Assessment of SLX4 Mutations in Hereditary Breast Cancers. PLoS One, 2013. 8(6): p. e66961.

61. Testa, J.R., et al., Germline BAP1 mutations predispose to malignant mesothelioma. Nature genetics, 2011. 43(10): p. 1022.

62. Wiesner, T., et al., Germline mutations in BAP1 predispose to melanocytic tumors. Nature genetics, 2011. 43(10): p. 1018.

63. Park, D., et al., Rare mutations in XRCC2 increase the risk of breast cancer. The American Journal of Human Genetics, 2012. 90(4): p. 734–739.

64. Vahteristo, P., et al., p53, CHK2, and CHK1 genes in Finnish families with Li-Fraumeni syndrome: further evidence of CHK2 in inherited cancer predisposition. Cancer research, 2001. 61(15): p. 5718–5722.

65. Majewski, I.J., et al., An α-E-catenin (CTNNA1) mutation in hereditary diffuse gastric cancer. The Journal of pathology, 2013. 229(4): p. 621–629.

66. Park, D.J., et al., Rare mutations in XRCC2 increase the risk of breast cancer. Am J Hum Genet, 2012. 90(4): p. 734–9.

67. Cybulski, C., et al., Germline RECQL mutations are associated with breast cancer susceptibility. Nature genetics, 2015. 47(6): p. 643.

68. Osher, D., et al., Mutation analysis of RAD51D in non-BRCA1/2 ovarian and breast cancer families. British journal of cancer, 2012. 106(8): p. 1460.

69. Varon, R., et al., Nibrin, a novel DNA double-strand break repair protein, is mutated in Nijmegen breakage syndrome. Cell, 1998. 93(3): p. 467–76.

70. Singh J, Thota N, Singh S et al (2018) Screening of over 1000 Indian patients with breast and/or ovarian cancer with a multi-gene panel: prevalence of BRCA1/2 and non-BRCA mutations. Breast Cancer Res Treat. 2018 Jul;170(1):189–196. doi:10.1007/s10549-018-4726-x.

71. Kim, H. and D.H. Choi, Distribution of BRCA1 and BRCA2 mutations in Asian patients with breast cancer. Journal of breast cancer, 2013. 16(4): p. 357–365.

72. Qamar, R., et al., Y-chromosomal DNA variation in Pakistan. The American Journal of Human Genetics, 2002. 70(5): p. 1107–1124.

73. Mehdi, S., et al., The Origins of Pakistani Populations, in Genomic Diversity. 1999, Springer. p. 83–90.

